# Mapping the Global Landscape of Task Shifting and Sharing: A Bibliographic Analysis from 1970 to 2022

**DOI:** 10.1101/2024.05.02.24306752

**Authors:** Shukanto Das, Liz Grant

## Abstract

Task shifting and sharing (TS/S) are strategies for redistributing healthcare services from more specialised to less-qualified providers. It aims to optimise service delivery, particularly in resource-constrained settings with workforce shortages. Our paper provides an overview of the global landscape of TS/S research, examining the geographic distribution, publication trends, variation in descriptors for TS/S and the disease focus of articles on TS/S. We searched five databases in October 2022, namely Medline, CINAHL Plus, Elsevier, Global Health and Google Scholar. Our bibliographic analysis included 2,072 articles related to TS/S. We extracted data on the countries where the studies were based, the terminology used to describe task redistribution, and the specific disease focus of the publications. The findings were then visualised and analysed to uncover trends and insights. The results revealed that TS/S research has been most extensively conducted in certain African and South Asian countries, particularly South Africa, India, Uganda, Kenya and Malawi. The terminology used to describe task redistribution varied, with “task shifting” being the most common term (66.0%), followed by “task sharing” (24.0%), “task delegation” (6.0%), and “task shifting and sharing” (3.9%). The disease focus of the publications was diverse, with HIV (n=450) and depression (n=375) being the most studied conditions, A major proportion of articles (42.5%) did not carry a disease focus, instead concentrated on broader health systems strengthening and policy issues. In conclusion, our study offers insights into the global landscape of TS/S research, highlighting the geographic disparities, terminology nuances and disease-specific applications. We believe our findings can inform future research and practice, including the need for standardisation of terminology, targeted implementation efforts, expansion of disease-specific applications and a focus on comprehensive systems strengthening. By addressing these considerations, stakeholders can optimise the impact of TS/S strategies and improve healthcare delivery and outcomes globally.

## Background

The global healthcare workforce is facing a serious crisis. At the 2017 Global Forum on Human Resources for Health (HRH), the World Health Organization (WHO) projected a need for approximately 40 million additional healthcare providers by 2030. It warned that the world could face a deficit of 18 million HRH, which would represent more than one in five of the 80 million healthcare providers required at that time [1,2]. The shortfall in HRH can be attributed to various factors. Challenges in attracting and retaining staff, particularly in remote regions [3,4], along with issues such as uneven distribution, high turnover rates and migration [5], insufficient investment in skills development and training [6], and a lack of both financial and non-financial incentives [1] are major contributors to the problem. Demographic and disease profile changes and weak health information systems, compound the crisis. The COVID-19 pandemic further highlighted deficiencies in healthcare systems worldwide, underscoring the urgent need for strategic planning, investment in infrastructure and support for HRH to become more resilience for future crises [7].

Indeed, addressing HRH shortages requires a multifaceted approach. Recruitment of providers can be increased via scholarships, loan forgiveness programs or other incentives to work in underserved areas [8]. Investing more in education and skill development, including in vocational training for healthcare professionals, can also help meet demands [6]. Improving working conditions, providing competitive salaries, benefits, opportunities for professional development and mentorship and addressing burnout can help retain staff [9–11]. Policy interventions, such as scope-of-practice reforms may enable providers such as nurse practitioners to undertake more duties independently and enhance the capabilities of existing HRH and alleviate shortages; the potential of which has been reviewed across several countries [12], including as the USA [13,14], India [15] and Israel [16].

Two approaches which align closely with the aforementioned strategies for addressing HRH shortages are task shifting and task sharing (TS/S). The WHO describes task shifting as the transfer or movement of tasks across professional hierarchies of HRH [17]. For example, the reassigning of particular services typically delivered by highly trained professionals, such as doctors, onto others who have lesser training in comparison, such as nurses or community health workers. Whereas task sharing is more collaborative in nature and can include a team of doctors, nurses or other allied professionals, leveraging each other’s skills [18–20]. Both strategies intend to optimise the use of available HRH and expand their scopes of practice. TS/S can be achieved by outlining responsibilities to be transferred, providing competency-focused training to new cadres and continuous monitoring and evaluating [17,18]. TS/S enhances the overall capacity of a system. By providing opportunities for professional growth and allowing HRH to expand their scope of practice, TS/S can increase job satisfaction and engagement and contribute to workforce retention. Moreover, by relieving overburdened staff and fostering a supportive environment, TS/S can mitigate burnout and improve work-life balance. TS/S has a long and successful history as a healthcare delivery strategy. Historical examples include the Chinese barefoot doctors [21] and Russian Feldshers [22], who delivered preventive care, diagnostics and emergency care to patients without access to specialist providers. TS/S has been used in surgery, obstetrics, anaesthesia and ophthalmology [23,24], as well as in caring for people living with human immunodeficiency virus [17,25], mental health services [26–28], managing hypertension, diabetes and obstructive lung diseases [29], offering family planning services [30] and screening for cervical cancer [31]. Task shifting was key in maintaining health services in the face of unprecedented patient surges during the pandemic. By delegating tasks to less-specialised providers, systems expanded capacities to respond to COVID-19, while continuing to address other pressing needs [32]. As healthcare needs continue to evolve post the pandemic, TS/S will continue to be crucial in creating sustainable healthcare workforces, strengthening resilience and ensuring delivery of improved quality services [32,33].

We are developing a new strategic implementation framework for TS/S as per the registered protocol cited here [34]. One of the early phases of this process was to review literature and map out evidences of TS/S from across all contexts and areas of healthcare. We were really interested to visualise the distribution of evidences or studies on TS/S on a global map and the year-wise distribution of these citations. We wanted to appreciate the variance in terminologies used to describe the process of TS/S between higher to lower cadres of HRH and examine which diseases or health conditions employed TS/S. To achieve this, we conducted a bibliographic analysis of published literature on TS/S. We chose to do bibliographic analysis as it allows parsing through thousands of citations at a time and examine relationships between research constituents, such as authors, countries or keywords [35]. Bibliographic analysis can be used to unpack trends, including emerging structures and themes, citation performances, collaboration patterns and priorities among scholars [36–38]. It is objective in nature, but can also be topped up with subjective thematic analysis, which can help meaningfully interpret literature [35]. For example, a bibliographic analysis on task shifting pertaining to nursing professionals from the year 2020, identified the most prolific scholars on the subject, connections within peers working on TS/S, as well as made a case for increased usage of digital technology in the facilitation of TS/S [39]. We sought to gain a broader overview of TS/S, considering all cadres of HRH, in order to place our research and the process of developing our implementation framework in the context of the larger landscape of health services research. In this paper, we report our findings from the bibliographic analysis on published literature on TS/S between the years 1970-2022.

## Methodology

We aimed to answer four questions: (A) What is the distribution of evidences or studies on TS/S as visualised on a world map? (B) What is the year-wise distribution of these evidences or studies? (C) What is the variance in terminologies used to describe TS/S between higher to lower cadres of HRH? (D) Which diseases or health conditions do these evidences or articles focus on? We used the following words and variations thereof to develop a search query: (“task shifting” OR “task sharing” OR “task shifting and task sharing” OR “task transfer” OR “task delegation” OR “TS/S”) AND (“healthcare” OR “health” OR “healthcare services”). We ran the search across five databases in October 2022, namely Medline, CINAHL Plus, Elsevier, Global Health and Google Scholar. The search was not restricted to a time period, aiming to find all articles published on TS/S from the beginning. Our search returned 5,754 citations. We imported these onto Covidence software [40] for screening. Duplications were removed. All records were manually screened by titles and abstracts. We included all articles related to the TS/S between higher to lower cadres of HRH. We included articles on both in-hospital and out-of-hospital settings, including all study designs, review articles and those from all geographical contexts. Articles covering any disease or health conditions were included. Only citations in English were included. PhD or masters dissertations were excluded. Irrelevant articles, such as studies relating to task switching in cognitive science and learning, were excluded. See **Fig1** for the flow diagram of search and selection process of citations. Detailed protocol is included as supplementary material **S1**. We used the BIBLIO checklist [41] to report our analysis (see supplementary material **S2**). SD performed the screening and LG supervised this to ensure accuracy and consistency. If there were doubts or discrepancies during screening, these were addressed via discussions. 2,072 citations were included in the final analysis. A spreadsheet containing titles, author list, full abstracts, publication years and indexing information was exported from Covidence. Supplementary file **S3** enlists all of these citations. We wrote a program in Python language and used Google Colaboratory [42] to run it and extract the following information from each citation: 1) Whether the study is based out of single or multiple countries and if so, a list of all these countries. A reference list containing country names and their alternate names was used to extract corresponding words from the spreadsheet. 2) What keywords or in-text terms are used to describe TS/S. A list of words such as “task shifting”, “task sharing”, “task shifting and sharing”, “task delegation”, “task transfer” and variations thereof was used as reference for extraction. 3) Which diseases or conditions do articles focus on. Disease names as per the WHO Global Health Estimates, which hosts the leading causes of death and disability globally [43], were used as reference. While most data were extracted successfully by the program, we found gaps in a few entries. These gaps were filled manually by SD. For instance, terminology data was not captured for 297 citations, so SD read full-texts of these papers and supplemented the information manually.

**Fig1:**
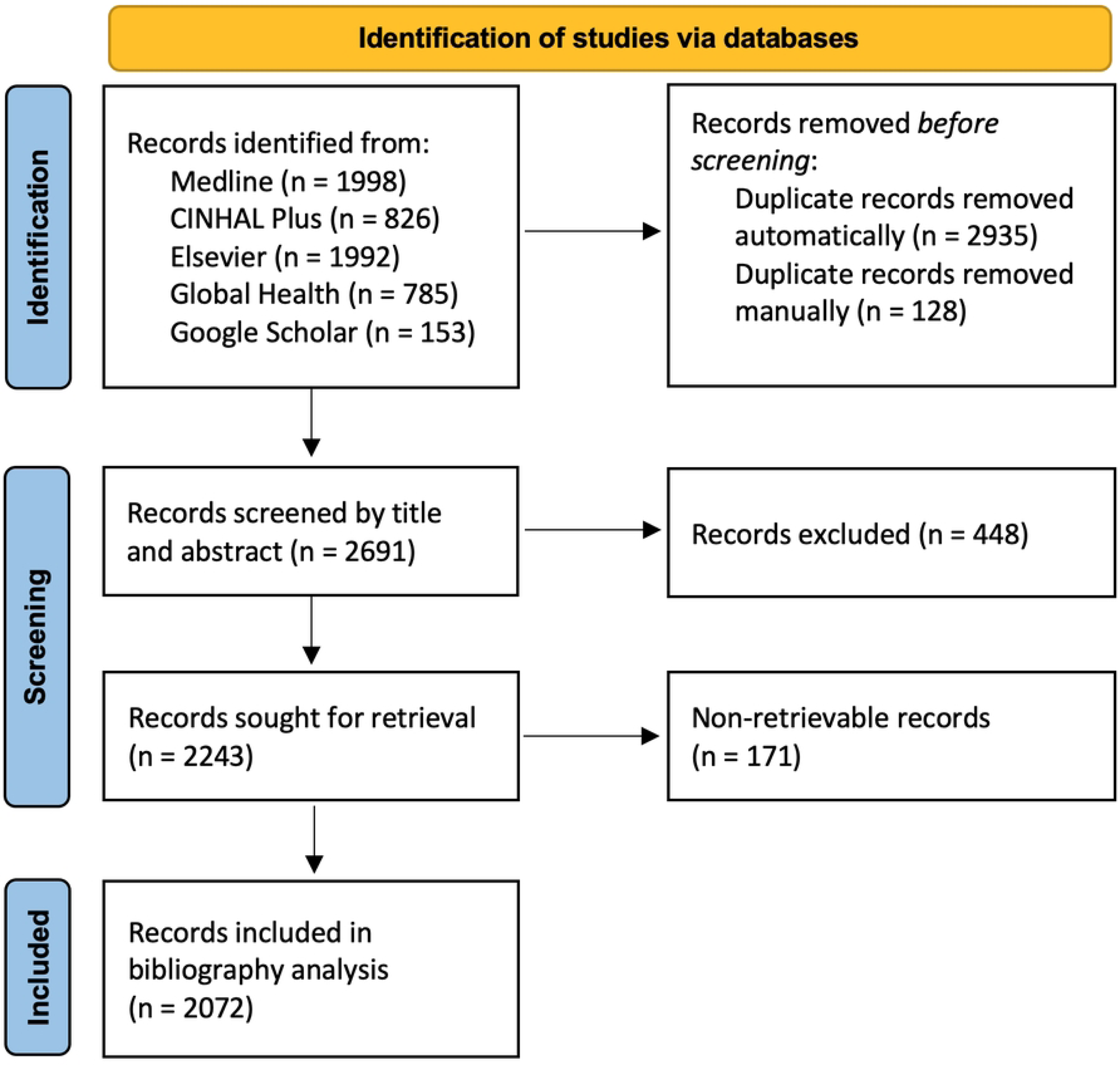
Flow diagram of search and selection process of citations.

## Results

Of the 2,072 articles in our analysis, titles or abstracts of 1,416 articles (68.3%) listed which countries these studies were based out of. 75.8% (n=1074) of these were based out of single countries. These were from across 99 countries; with highest number of publications based on TS/S in South Africa (n=183), India (n=115), Uganda (n=77), Kenya (n=74) and Nigeria (n=58). Factoring in the other 342 multi-country studies, we noted that TS/S has been most extensively studied in South Africa (n=201), followed by India (n=130), Uganda (n=109), Kenya (n=106) and Malawi (n=86). **Fig2** shows the geographic distribution of publications on TS/S on the world map. Note that only papers containing single-country studies are considered in this visualisation. Furthermore, we used World Bank data [44] to visualise the distribution of these countries as per their income levels (See **Fig3**). Low middle-income countries (n=30), including India, Kenya and Nigeria, have published the most on TS/S. USA, Canada, UK and other high-income countries (n=29) follow at close second. Upper middle-income countries (n=20), including South Africa, Brazil, China and low-income countries (n=20) such as Uganda, Malawi and Ethiopia, have published on TS/S equally. Since our search was not limited to a time period, our analysis reported articles on TS/s from as early as the year 1970. **Fig4** shows the trends of publications on TS/S from 1970 up until 2022 October. We noted a rapid rise in publications since 2006–2008.

**Fig2:**
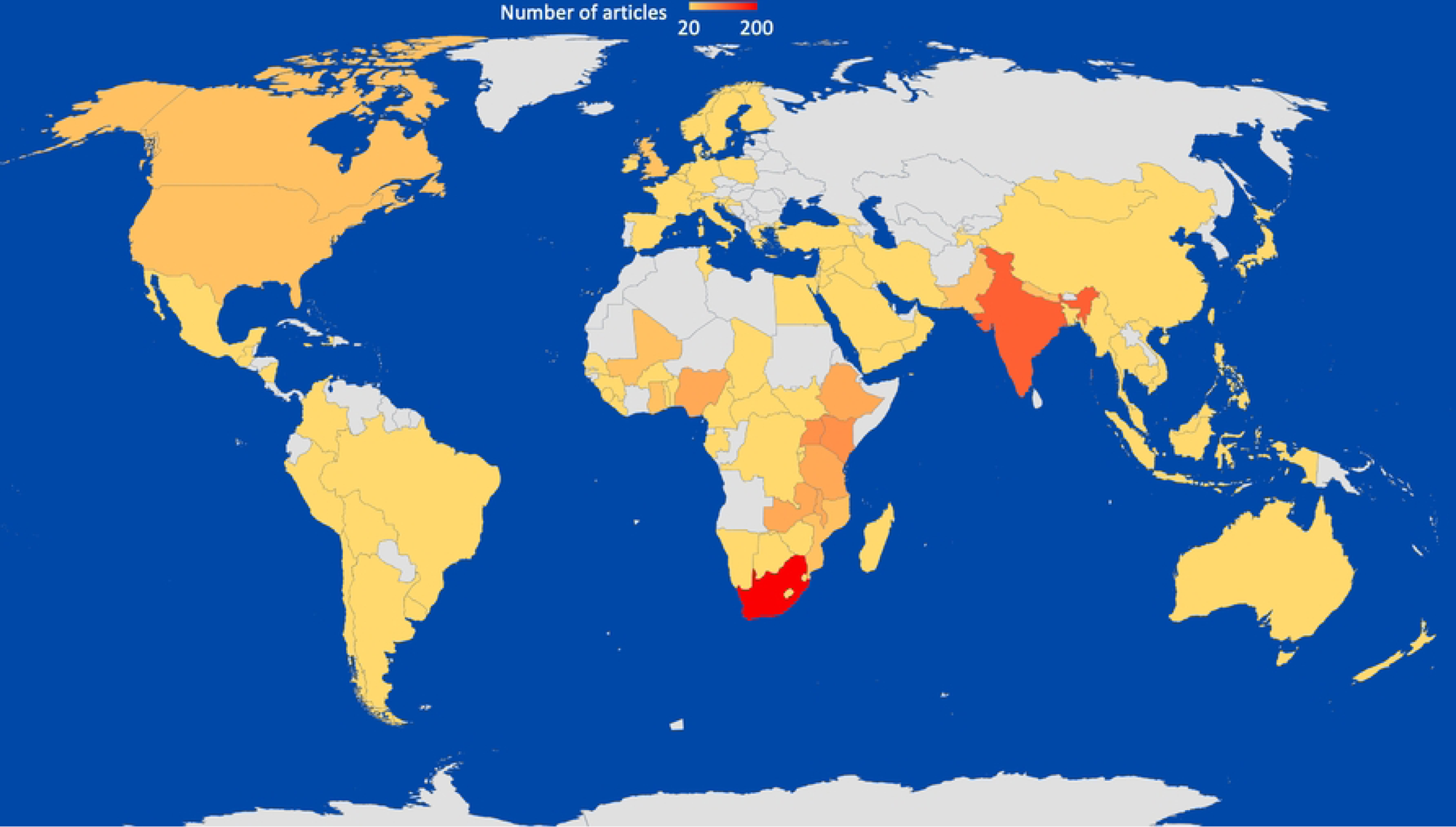
Global distribution of publications on task shifting and sharing (1970-2022). An increasing number of publications is represented by the gradient from light yellow (low) to bright red (high). Countries shown in white indicate no papers in our analysis.

**Fig3:**
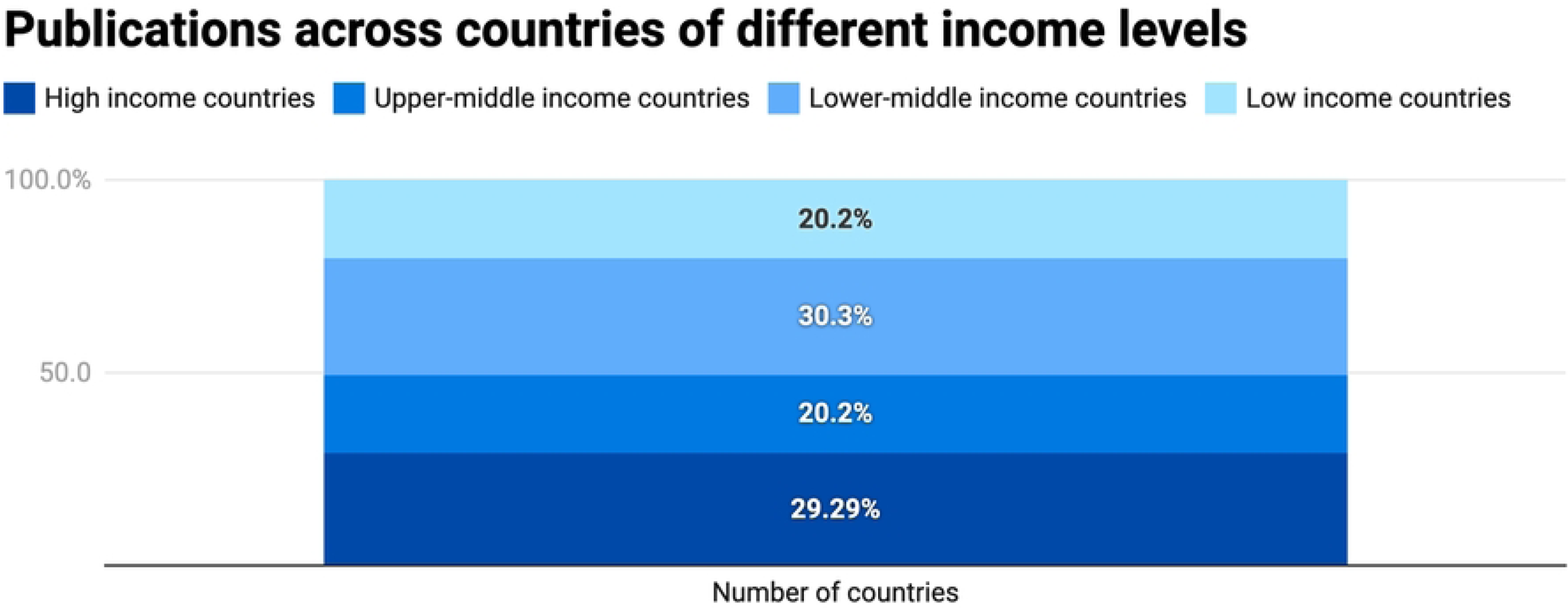
Distribution of countries with publications on TS/S between 1970–2022, categorised by their income-level profiles

**Fig4:**
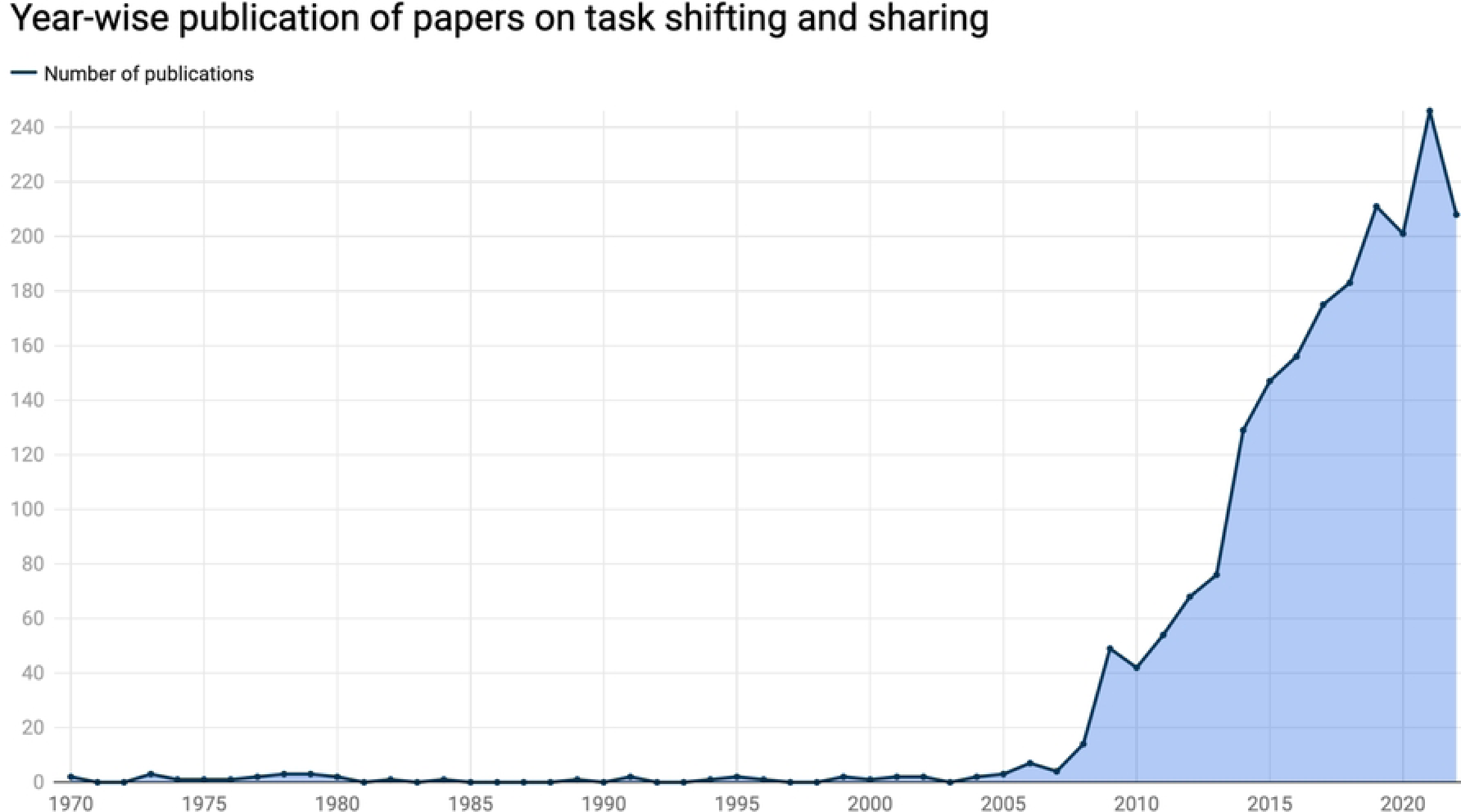
Publication trends of papers on TS/S between years 1970–2022

Articles varied in how they describe the movement of tasks from higher to lower cadres of HRH. 1,369 publications used the words “task shifting” (66.0%), 492 articles used “task sharing” (24.0%), 117 articles used “task delegation” (6.0%), 81 articles expressed it as “task shifting and sharing” (3.9%) and the other 13 articles used “task transfer”. **Fig5** depicts this distribution. Furthermore, with regards to the disease focus of the articles in the study, only 1,192 (57.5%) citations report a specific disease focus. These are spread across a total of 37 diseases categories, as per the Global Health Estimates classification [43]. HIV (n=450) and depression (n=375) are the most studied diseases with respect to TS/S; followed by publications on diabetes (n=69), tuberculosis (n=65), anxiety (n=40), maternal health (n=32), hepatitis (n=23) and more. **Fig6** depicts this distribution. While the other 882 articles did not carry disease names in their abstracts or titles, they focus on issues such as overall health systems strengthening, resource and services organisation, health policy and health economics.

**Fig5:**
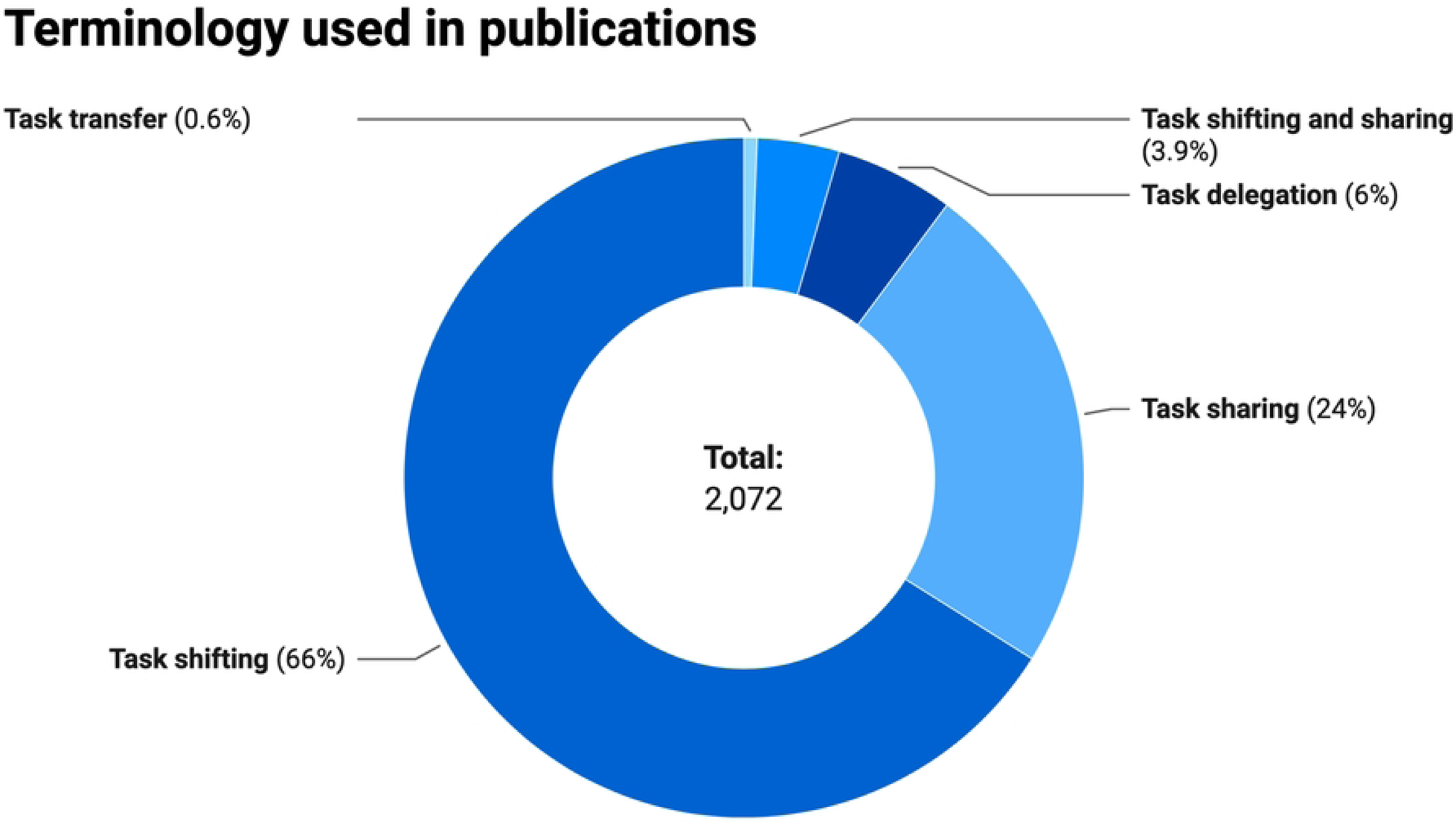
Variation of terminology used to describe task shifting and sharing used in publication between 1970–2022

**Fig6:**
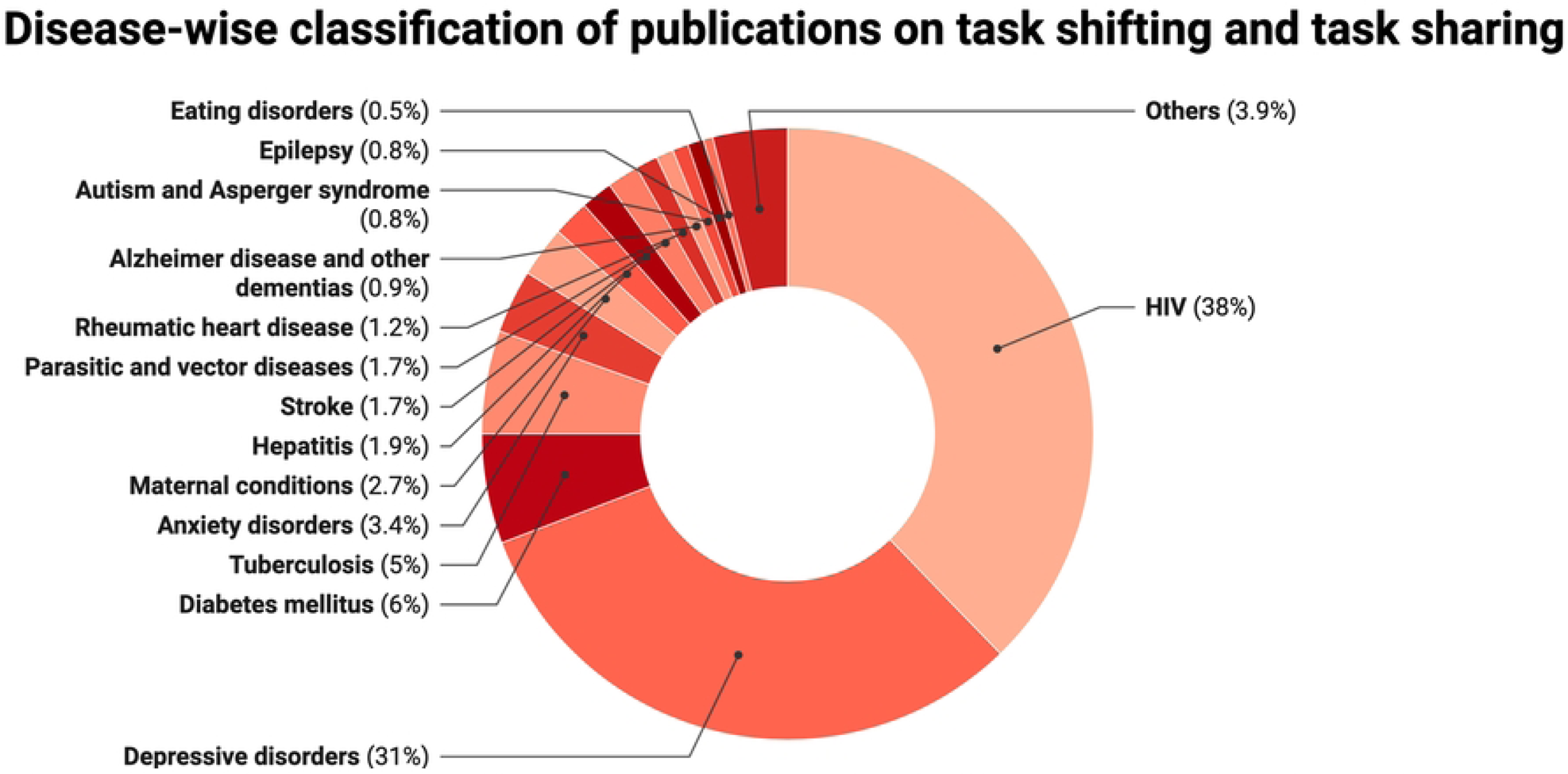
Classification of papers on task shifting and sharing between 1970–2022 as per their disease focus

## Discussion

We present the findings grouped under the four questions discussed previously in the methodology section.

### Global distribution of research on task shifting and sharing

Our analysis provides insights into the geographic distribution of research on TS/S. Findings indicate that TS/S has been most extensively studied in certain African and South Asian regions, particularly South Africa, India, Uganda, Kenya and Malawi. The high volume of publications on TS/S originating from these nations suggests that the practice of TS/S has been a focus of research or practice in these low-and middle-income regions. This aligns with the broader literature, which has also remarked TS/S as an key strategy for expanding access to services in resource-constrained settings, where HRH shortages are most acute. Interestingly, our results show that low and lower-middle income countries, such as India, Kenya and Nigeria, have published the most on TS/S, followed closely by high-income countries like the USA, Canada and the UK. This indicates that TS/S is a topic of global interest and relevance, with research being conducted across a range of economic contexts. The finding that upper-middle income countries, including South Africa, and low-income countries, such as Uganda and Malawi, have published on TS/S to a similar extent is also noteworthy. This suggests that the challenges and opportunities around TS/S may transcend strict income classifications, and lessons learned in one setting may be applicable to others. It would be crucial to explore the particular drivers and facilitators that have enabled TS/S research and implementation efforts in low- and middle-income settings. Factors like healthcare workforce gaps, disease burden, health infrastructure and policy environments are likely key considerations. Analysing these could provide insights for replicating and scaling successful TS/S models in similar resource-limited regions. Conversely, the strong research presence from high-income countries also merits exploration. While TS/S may not be as critical a strategy in these resource-abundant settings, the involvement of these nations suggests that TS/S holds relevance and applicability across their contexts too. Understanding the use cases and outcomes of TS/S initiatives in high-income countries could yield valuable lessons for optimising the strategy globally.

### Publications trends over the past 50 years

The temporal aspect, showing publications on TS/S dating back to the 1970s, showcases the interest in TS/S garnered over time. Articles by Bates, advocating expansion of nursing and physician assistant roles through delegation [45], and by Yankauer *et al*, on the delegation of ambulatory tasks by pediatricians to increase productivity [46], are first examples of work on TS/S from our analysis. One of the initial increases in the publication volume is around the year 1978. This could be associated with the Declaration of Alma-Ata [47], as has also been suggested by Benton *et al* in their analysis [39]. The Alma-Ata Declaration encouraged community involvement and resource optimisation in primary health care. It recommended providers and stakeholders of primary care to actively coordinate and collaborate in order to make services delivery more efficient, which after all are key principles for TS/S. But we truly see a rapid rise in publications since 2006–2008, which can be associated with the launch of two global reports. Firstly, the WHO ‘World Health Report: Working together for health’ of 2006, which estimated the shortages of HRH in 57 countries to be over 2.4 million and proposed strategies to tackle this crisis over the next decade through delegation and task shifting [48]. And then, the report of 2008 by WHO, the Joint United Nations Programme on HIV and AIDS (UNAIDS) and the USA’s President’s Emergency Plan for AIDS Relief (PEPFAR) titled ‘Task shifting: Rational redistribution of tasks among health workforce teams – Global recommendations and guidelines’ which presented 22 evidence-based recommendations to adopt task shifting as national strategies and reorganise HRH and healthcare and financial resources [17].

### Variance in terminology used to describe task shifting and sharing

Majority of the published articles using the term “task shifting” to articulate TS/S suggests that it’s the most commonly recognised and accepted descriptor for the practice of moving healthcare tasks from higher-level to lower-level cadres. The use of other terms like “task sharing”, “task delegation” and “task shifting and sharing” indicates that there is some variation in how the process is articulated. This variance reflects the multifaceted nature of redistributing tasks among different cadres of HRH and may have stemmed from differences in regional practices, policy frameworks and protocolisation of task redistribution in systems. This variability however, can cause confusion, especially when researchers are synthesising new findings or when policies are being developed or implemented. Understanding these variations can help in standardising terminology and establishing clearer communication and collaboration. Researchers, healthcare providers, policy makers and other stakeholders must therefore concur on meanings and accepted terminology while studying and reporting TS/S.

### Disease profile of publications on task shifting and sharing

Interestingly, only 57.5% of the articles in our analysis reported a specific disease focus. The distribution of disease focus in TS/S research, with HIV and depression emerging as the most studied diseases, sheds light on health priorities that drove research and implementation efforts on TS/S. The prominence of HIV and depression reminds us of their huge burdens and the potential impact that TS/S can have in addressing these complex challenges. The breadth of disease categories covered, spanning 37 different conditions, also speaks of the versatility of TS/S as a tool. However, the relatively small number of publications on certain high-burden conditions like tuberculosis (n=65) and maternal health (n=32) indicates that there may be opportunities to further explore the potential of TS/S in these domains. A major proportion of publications from our sample did not specify a disease focus, but instead concentrated on broader health systems strengthening, resource organisation and health policy aspects. This reflects a recognition of the interconnectedness of various system components and the need for comprehensive approaches in service delivery assisted by TS/S, that go beyond disease-specific interventions.

### Limitations and implications of future research

Our methodology presents a limitation that the data on country and disease names was extracted solely from the titles and abstracts of the publications. The underlying assumption was that if articles were based in specific countries or had a disease focus, this information would be explicitly stated in the titles or abstracts. However, it is possible that some articles may have mentioned these details only in the main text, which would not have been captured by our method. This could result in an incomplete representation of the geographic distribution and disease focus of the TS/S research. Future studies might want to address this by incorporating more comprehensive data extraction methods, such as full-text analysis. This however would be difficult, given the volume of publications on TS/S.

Despite this limitation, the insights gained from this study can inform future research and practice. Given the wide geographic distribution of TS/S, future studies should explore drivers, facilitators and barriers to TS/S adoption in different healthcare systems, including low- and middle-income countries and high-income settings. Variations in terminology used to describe TS/S also calls for standardisation. Establishing definitions and frameworks will enhance communication, collaboration and comparability across settings. It will also streamline discussions, knowledge sharing and implementation of TS/S. The focus on TS/S in diseases such as HIV, depression, diabetes and tuberculosis indicates the potential impact of TS/S on other health priorities as well. More studies exploring effectiveness in different disease contexts, identifying best practices and assessing scalability and sustainability across settings would be helpful. Future studies should also delve into system-wide implications of integrating TS/S and assess long-term sustainability and equity.

## Conclusion

Our bibliographic analysis has examined a large dataset of published research articles between 1970-2022 and thrown light on the global landscape of TS/S research, highlighting trends and priorities in the process. The unique contribution of this study is that it visualises the geographic distribution of TS/S research, revealing the prominence of certain low- and middle-income countries like South Africa, India and Uganda, as hotspots for research on task redistribution. Additionally, the study has discovered the variance in terminologies used to describe TS/S practices and the varying disease focuses of research publications, providing a nuanced understanding of the multifaceted nature of TS/S strategies. Our findings offer insights for policymakers, researchers and healthcare practitioners seeking to understand and optimise the implementation of TS/S.

## Data Availability

All relevant data supporting the findings are within the paper and its Supporting information files.

## Author contributions

Both authors conceptualised the study together. SD developed the study protocol, wrote the codes, analysed data and drafted the manuscript. LG reviewed the data analysis and the manuscript. Authors received no specific funding for this work. Authors declare no conflict of interest.

## Supplementary information

**S1: Protocol used to conduct literature search and data analysis S2: BIBLIO checklist for reporting data**

**S3: Spreadsheet enlisting data of citations included in the study**

## References

1. Liu JX, Goryakin Y, Maeda A, Bruckner T, Scheffler R. Global Health Workforce Labor Market Projections for 2030. Hum Resour Health. 2017;15. doi:10.1186/s12960-017-0187-2. [cited 24 April 2024]

2. Bosanquet N. Human: solving the global workforce crisis in healthcare. British Journal of Healthcare Management. 2019;25. doi:10.12968/bjhc.2019.25.5.206. [cited 24 April 2024]

3. Khalil M, Alameddine M. Recruitment and retention strategies, policies, and their barriers: A narrative review in the Eastern Mediterranean Region. Health Science Reports. 2020. doi:10.1002/hsr2.192. [cited 24 April 2024]

4. Jaeger FN, Bechir M, Harouna M, Moto DD, Utzinger J. Challenges and opportunities for healthcare workers in a rural district of Chad. BMC Health Serv Res. 2018;18. doi:10.1186/s12913-017-2799-6. [cited 24 April 2024]

5. Kamarulzaman A, Ramnarayan K, Mocumbi AO. Plugging the medical brain drain. The Lancet. 2022. doi:10.1016/S0140-6736(22)02087-6. [cited 25 April 2024]

6. Cancedda C, Farmer PE, Kerry V, Nuthulaganti T, Scott KW, Goosby E, et al. Maximizing the Impact of Training Initiatives for Health Professionals in Low-Income Countries: Frameworks, Challenges, and Best Practices. PLoS Med. 2015;12. doi:10.1371/journal.pmed.1001840. [cited 25 April 2024]

7. Rogers HL. The organisation of resilient health and social care following the COVID-19 pandemic – A critical review. Eur J Public Health. 2021;31. doi:10.1093/eurpub/ckab164.323. [cited 25 April 2024]

8. Schwartz MR, Patterson DG, McCarty RL. State Incentive Programs that Encourage Allied Health Professionals to Provide Care for Rural and Underserved Populations. 2019 Dec. [cited 25 April 2024]

9. Razai MS, Kooner P, Majeed A. Strategies and Interventions to Improve Healthcare Professionals’ Well-Being and Reduce Burnout. Journal of Primary Care and Community Health. 2023. doi:10.1177/21501319231178641. [cited 26 April 2024]

10. Miloslavsky EM, Bolster MB. Addressing the rheumatology workforce shortage: A multifaceted approach. Seminars in Arthritis and Rheumatism. 2020. doi:10.1016/j.semarthrit.2020.05.009. [cited 26 April 2024]

11. Hookmani AA, Lalani N, Sultan N, Zubairi A, Hussain A, Hasan BS, et al. Development of an on-job mentorship programme to improve nursing experience for enhanced patient experience of compassionate care. BMC Nurs. 2021;20. doi:10.1186/s12912-021-00682-4. [cited 26 April 2024]

12. Htay M, Whitehead D. The effectiveness of the role of advanced nurse practitioners compared to physician-led or usual care: A systematic review. International Journal of Nursing Studies Advances. 2021. doi:10.1016/j.ijnsa.2021.100034. [cited 26 April 2024]

13. Iglehart JK. Expanding the Role of Advanced Nurse Practitioners — Risks and Rewards. New England Journal of Medicine. 2013;368. doi:10.1056/nejmhpr1301084. [cited 26 April 2024]

14. Poghosyan L, Ghaffari A, Liu J, Jin H, Martsolf G. State policy change and organizational response: Expansion of nurse practitioner scope of practice regulations in New York State. Nurs Outlook. 2021;69. doi:10.1016/j.outlook.2020.08.007. [cited 26 April 2024]

15. Nanda L, Anilkumar A. Role of nurse practitioners within health system in India: A case of untapped potential. J Family Med Prim Care. 2021;10. doi:10.4103/jfmpc.jfmpc_2283_20 [cited 26 April 2024]

16. Maier CB, Aiken LH. Expanding clinical roles for nurses to realign the global health workforce with population needs: A commentary. Israel Journal of Health Policy Research. 2016. doi:10.1186/s13584-016-0079-2. [cited 26 April 2024]

17. World Health Organization. Task Shifting: Global Recommendations and Guidelines. World Health Organization. 2008 [cited 20 Oct 2022]. doi:10.1080/17441692.2011.552067. [cited 27 April 2024]

18. Centre for Disease Control and Prevention. Sharing and Shifting Tasks to Maintain Essential Healthcare During COVID-19 in Low Resource, non-US settings. In: National Center for Immunization and Respiratory Diseases (NCIRD), Division of Viral Diseases. 17 Jun 2022. [cited 27 April 2024]

19. Lunsford S, Broughton E, Fatta K. Task Shifting/Sharing for HIV Services in 26 PEPFAR-supported Countries: A Qualitative Assessment - Research and Evaluation Report. 2019. Available: https://pdf.usaid.gov/pdf_docs/PA00WGCS.pdf. [cited 27 April 2024]

20. Tsui S, Denison JA, Kennedy CE, Chang LW, Koole O, Torpey K, et al. Identifying models of HIV care and treatment service delivery in Tanzania, Uganda, and Zambia using cluster analysis and Delphi survey. BMC Health Serv Res. 2017;17(1): 811. doi:10.1186/s12913-017-2772-4. [cited 27 April 2024]

21. Gross M. Between Party, People, and Profession: The Many Faces of the “Doctor” during the Cultural Revolution. Med Hist. 2018;62(3): 333–359. doi:10.1017/mdh.2018.23. [cited 27 April 2024]

22. Sidel VW. Feldshers and Feldsherism. New England Journal of Medicine. 1968;278(18): 987–92. doi:10.1056/nejm196804252781705. [cited 25 April 2024]

23. Vaz F, Bergström S, Vaz MDL, Langa J, Bugalho A. Training medical assistants for surgery. Bull World Health Organ. 1999;77(8): 688–91. Available: https://apps.who.int/iris/handle/10665/267901. [cited 27 April 2024]

24. Mullan F, Frehywot S. Non-physician clinicians in 47 sub-Saharan African countries. Lancet. 2007;370(9605): 2158–63. doi:10.1016/S0140-6736(07)60785-5. [cited 27 April 2024]

25. Zachariah R, Ford N, Philips M, S. Lynch, Massaquoi M, Janssens V, et al. Task shifting in HIV/AIDS: opportunities, challenges and proposed actions for sub-Saharan Africa. Transactions of the Royal Society of Tropical Medicine and Hygiene. 2009. pp. 549–58. doi:10.1016/j.trstmh.2008.09.019. [cited 27 April 2024]

26. Kola L, Kohrt BA, Hanlon C, Naslund JA, Sikander S, Balaji M, et al. COVID-19 mental health impact and responses in low-income and middle-income countries: reimagining global mental health. Lancet Psychiatry. 2021;8(6): 535–550. doi:10.1016/S2215-0366(21)00025-0. [cited 28 April 2024]

27. Javed A, Lee C, Zakaria H, Buenaventura RD, Cetkovich-Bakmas M, Duailibi K, et al. Reducing the stigma of mental health disorders with a focus on low- and middle-income countries. Asian J Psychiatr. 2021;58: 102601. doi:10.1016/j.ajp.2021.102601. [cited 28 April 2024]

28. Zhang Y, Lange K. Coronavirus disease 2019 (COVID-19) and global mental health. Global Health Journal. 2021;5(1): 31–36. doi:10.1016/j.glohj.2021.02.004. [cited 27 April 2024]

29. Mobula LM, Heller DJ, Commodore-Mensah Y, Walker Harris V, Cooper LA. Protecting the vulnerable during COVID-19: Treating and preventing chronic disease disparities. Gates Open Res. 2020;4: 125. doi:10.12688/gatesopenres.13181.1. [cited 27 April 2024]

30. Johnson SA, Kaggwa MN, Lathrop EVA. How It Started, and How It’s Going: Global Family Planning Programs. Clin Obstet Gynecol. 2021;64(3): 422–434. doi:10.1097/GRF.0000000000000625. [cited 27 April 2024]

31. Steben M, Norris T, Rosberger Z, HPV Global Action. COVID-19 Won’t Be the Last (Or Worst) Pandemic: It’s Time to Build Resilience Into Our Cervical Cancer Elimination Goals. Journal of Obstetrics and Gynaecology Canada. 2020;42(10): 1195–1196. doi:10.1016/j.jogc.2020.08.006. [cited 29 April 2024]

32. Das S, Grant L, Fernandes G. Task shifting healthcare services in the post-COVID world: A scoping review. PLOS Global Public Health. 2023;3: e0001712. doi:10.1371/journal.pgph.0001712. [cited 27 April 2024]

33. Expert Panel on effective ways of investing in Health (EXPH). Task shifting and health system design. 2019 Jun. Available: https://health.ec.europa.eu/system/files/2019-11/023_taskshifting_en_0.pdf. [cited 29 April 2024]

34. Das S, Grant L, Weller D. Mixed-methods study to validate and refine the “Strategic Healthcare Implementation Framework for Task Shifting, Sharing and Resource Enhancement” (SHIFT-SHARE). protocols.io. 2023 [cited 2 Aug 2023]. doi:10.17504/protocols.io.36wgq35zylk5/v1. [cited 27 April 2024]

35. Donthu N, Kumar S, Mukherjee D, Pandey N, Lim WM. How to conduct a bibliometric analysis: An overview and guidelines. J Bus Res. 2021;133. doi:10.1016/j.jbusres.2021.04.070. [cited 29 April 2024]

36. Donthu N, Kumar S, Pandey N, Lim WM. Research Constituents, Intellectual Structure, and Collaboration Patterns in Journal of International Marketing: An Analytical Retrospective. Journal of International Marketing. 2021;29. doi:10.1177/1069031X211004234. [cited 29 April 2024]

37. Ellegaard O, Wallin JA. The bibliometric analysis of scholarly production: How great is the impact? Scientometrics. 2015;105. doi:10.1007/s11192-015-1645-z. [cited 29 April 2024]

38. Wallin JA. Bibliometric methods: Pitfalls and possibilities. Basic and Clinical Pharmacology and Toxicology. 2005. doi:10.1111/j.1742-7843.2005.pto_139.x. [cited 30 April 2024]

39. Benton DC, Ferguson SL, Holloway A. Task Shifting: A High-Level Analysis of Scholarship. J Nurs Regul. 2020;11. doi:10.1016/S2155-8256(20)30104-6. [cited 29 April 2024]

40. Veritas Health Innovation. Covidence systematic review software. Melbourne Australia. Melbourne, Australia; Available: www.covidence.org. [cited 28 April 2024]

41. Montazeri A, Mohammadi S, M. Hesari P, Ghaemi M, Riazi H, Sheikhi-Mobarakeh Z. Preliminary guideline for reporting bibliometric reviews of the biomedical literature (BIBLIO): a minimum requirements. Syst Rev. 2023;12. doi:10.1186/s13643-023-02410-2. [cited 29 April 2024]

42. Bisong E. Google Colaboratory. Building Machine Learning and Deep Learning Models on Google Cloud Platform. 2019. doi:10.1007/978-1-4842-4470-8_7. [cited 28 April 2024]

43. World Health Organization. Global health estimates: Leading causes of death. 2023. Available: https://www.who.int/data/gho/data/themes/mortality-and-global-health-estimates/ghe-leading-causes-of-death. [cited 29 April 2024]

44. The World Bank. World Bank list of economies (June 2020). World Bank list of economies. 2020. [cited 28 April 2024]

45. Bates B. Doctor and nurse: Changing roles and relations. Nurs Manage. 1970;1. doi:10.1097/00006247-197010000-00004. [cited 28 April 2024]

46. Yankauer A, Connelly JP, Feldman JJ. Physician productivity in the delivery of ambulatory care: Some findings from a survey of pediatricians. Med Care. 1970;8. doi:10.1097/00005650-197001000-00005. [cited 29 April 2024]

47. World Health Organization. Declaration of Alma-Ata. In: https://iris.who.int/bitstream/handle/10665/347879/WHO-EURO-1978-3938-43697-61471-eng.pdf. 1978. [cited 30 April 2024]

48. World Health Organization. Working together for Health: World Health Report 2006. World Health Organization. Geneva; 2006. Available: https://apps.who.int/iris/handle/10665/43432. [cited 29 April 2024]

